# Cell-Type-Resolved Transcriptional Remodelling in Parkinson’s Disease Substantia Nigra: An Integrated Framework Implicates NPAS3 and BNC2 Regulatory Subnetworks in Dopaminergic Neurons and Glial Subpopulations

**DOI:** 10.64898/2026.06.04.26354575

**Authors:** Sana Noor, Fazila Zahoor

## Abstract

**Background:** Parkinson’s disease (PD) is a second most common progressive neurological disorder that is pathologically characterized by the loss of dopaminergic neurons within the substantia nigra (SN). However, disease progression probably involves coordinated changes across both neuronal and glial cell populations. Although single-nucleus RNA-seq resolved cell-type-specific transcriptional profiling, differential expression and regulatory interpretation are commonly reported separately, however, they may limit the mechanistic prioritization to uncover novel therapeutic targets.

**Methods:** Here, we performed sample-aware pseudobulk framework analysis on single-nucleus transcriptomes obtained SN of PD and control donors. Cell-type-specific differential expression for PD vs. control was identified using edgeR quasi-likelihood modeling (FDR < 0.05; |log2FC| > 0.5). Further, to quantify disease-specific remodelling, we computed one-vs-rest cell-type specificity scores in each condition and defined delta-specificity as the PD–control shift. We further prioritized the gene-set for dopaminergic neurons and microglia based on edge R significance and delta-specificity shifts, followed by upstream regulatory assessment using transcription factor enrichment and subnetwork visualization using ChEA-KG. Moreover, we used Cellchat to identify altered cell-cell communication networks to infer differences between both conditions.

**Results:** Dopaminergic neurons demonstrated upregulation of neuronal-state remodeling transcriptional programs related gene sets in PD group, including receptor signaling and contact/guidance pathways (e.g., CHRM3, ROBO1, PLXNA4, UNC5D, EFNA5), neuronal excitability homeostatsis, RNA components, cellular traffickings and proteostasis, suggesting coordinated remodeling in surviving neuronal population. Microglia exhibited a compact PD-associated signature enriched for regulatory and activation state-related genes. TF networks analysis revealed distinct regulatory subnetwork in each population,including BNC2-centered network in microglia and an NPAS3-centered network in dopaminergic neurons with embedded ZNF804A and chromatin-associated components.

**Conclusions:** In summary, integrating pseudobulk, delta-specificity scoring and TF-network enrichment analysis provides coherent dopaminergic and microglial programs in PD substantia nigra. This framework prioritizes cell-type-specific potential candidate mechanisms for downstream validation. The inferred regulatory networks and interactions are hypothesis generating and need orthogonal validation, such as spatial or proteomics approaches and independent cohorts.

## 1. Introduction

Parkinson’s disease (PD) is the second most frequently occurring chronic neurodegenerative disorder after Alzheimer, with a multifactorial etiology and affects over 10 millions of people worldwide (Ben-Shlomo et al. 2024; Dong-Chen et al. 2023; Li et al. 2025). It is characterized by progressive loss of dopamine-producing neurons in substantia nigra (SN) and formation of alpha synuclein (α-syn) aggregates into intracellular inclusions, forming in Lewy bodies (LBs) in the soma and Lewy neurites (LNs) in dendrites and axons (Calabresi et al. 2023; Magalhães and Lashuel 2022; Spillantini et al. 1997). It represents a range of clinical features that includes both motor and non-motor symptoms (Dong-Chen et al. 2023). Motor symptoms primarily comprise rigidity, postural instability, bradykinesia and tremor (Chaudhary, et al. 2025). However, PD patients also have a broad spectrum of non-motor symptoms, such as sleep disturbance, depression, cognitive decline, hyposmia and autonomic urinary dysfunctional (Armstrong and Okun 2020). In addition to neuropathological hallmarks of PD, other molecular and cellular mechanism disruption leads to neuronal homeostasis. Activation of microglia (Calabresi et al. 2023; Schetters et al. 2018), dysregulated innate and adaptive immune responses (DeMaio et al. 2022), oxidative stress, (Chaudhary et al. 2025) and mitochondrial dysfunctions (Calabresi et al. 2023).

Despite the advances in PD research, our understanding of the exact molecular mechanism beyond PD remains inadequate to uncover the exact therapeutic effect and its associated signature. However, development of multi-omics technologies specifically single cell sequencing (ScRNA-seq) and single nuclei sequencing (SnRNA-seq) as well as advance computational analysis has substantially expanded our understanding of complex diseases such as PD by providing molecular-level characterization of distinct neuronal and non-neuronal heterogeneous cellular populations and how molecular pathways are altered in these specific cell type in disease condition (Ma & Lim 2021). Both ScRNA-seq and SnRNA-seq have identified significant heterogeneity among dopaminergic neuron subpopulations, identification of markers such as Grp and Neurod6 that helps to distinguish between vulnerable and resilient neuronal types.

Furthermore, these advanced approaches uncovered how genetic mutations LRRK2-G2019S PARKIN, and PINK LRRK2-G2019S can enhance neuronal differentiation and death (Mirzac et al. 2025; Yang et al. 2024). However, high resolution profiling of immune cells including peripheral immune cell (Moquin-Beaudry et al. 2025), microglia (Tichauer et al. 2023) and CNS-associated macrophages (Yang et al. 2024) have revealed cell-type and cellular state-specific transcriptional changes which may lead to neuroinflammation, α-syn pathology and blood brain barrier dysfunction in PD patients (Dhanwani et al. 2022; Yang et al. 2024)

PD is defined by degeneration of dopaminergic neurons in the SN, but the molecular basis of this vulnerability is not purely neuron-intrinsic. Increasing evidence indicates that glial state changes (especially microglia) (Martirosyan et al. 2024) and altered neuron–glia communication contribute to SN dysfunction and progression (Huang et al. 2022; Yi et al. 2022). Recent single-nucleus/single-cell transcriptomic profiling of the human substantia nigra has provided cell-type-resolved maps of PD-associated molecular programs, including glial state remodeling and neuronal vulnerability signatures (Martirosyan et al. 2024; Wang et al. 2024). However, many studies emphasize differential expression and cell-state shifts as separate outputs, and only a subset explicitly interrogates how disease-associated transcriptional programs relate to intercellular communication architecture. To address this gap, we integrate three complementary views, sample-aware pseudobulk DE (edgeR), cell-type identity remodeling (Δ-specificity), neuron-glial remodeling and regulatory program shifts, and inferred cell–cell communication (CellChat), to provide a unified, cell-type-resolved map of PD remodeling in substantia nigra.

## 2. Methodology

### SnRNA-Seq Data Download and Pre-Processing

We downloaded the publicly available single-nucleus transcriptomics dataset from the GEO database (https://www.ncbi.nlm.nih.gov/geo/) under accession number GSE243639. The dataset contains 29 post-mortem substantia nigra pars compacta (SNpc) brain samples, including 15 Parkinson’s disease samples and 14 controls (Martirosyan et al. 2024). To process the raw expression matrix, Seurat v5.3.1.1 (Satija Lab, USA) was used. For each control and diseased SnRNA-seq data, a Seurat object was created using the function c*reateSeuratObject,* and all the objects were merged for further processing. Low-quality nuclei were filtered out by applying the following thresholds: cells with more than 200 RNA features, fewer than 2000 features, or fewer than 5% mitochondrial genes were included.

### SnRNA -Seq, Normalization, Scaling, Dimension Reduction, and Clustering

After quality control, the dataset was normalized using the LogNormalize method with a scale factor of 10,000 implemented in Seurat. We applied the variance stabilizing transformation (VST) method to identify highly variable genes (HVGs), and the top 2000 HVGs were identified, scaled, and then centered. Next, dimensionality reduction was done by principal component analysis on the top HVGs. The FindNeighbors function was used to calculate cell-cell similarities, based on selected principal components, followed by clustering using the Louvain algorithm with a resolution of 0.5. Finally, Uniform Manifold Approximation and Projection (UMAP) and t-distributed stochastic neighbor embedding (t-SNE) were employed to reduce non-linear dimensionality and visualize cell clusters.

### Clusters Annotation and DEGs Analysis

ScType (Ianevski, Giri, and Aittokallio 2022) was employed to annotate the identified clusters based on predefined marker gene sets. To identify brain-specific gene sets, the “gene_sets_prepare” function was used to access both positive and negative markers from the ScType database. scType scores were calculated based on the expression of these markers, and cluster-level aggregation of these scores identified cluster-specific cell types. Moreover, to minimize false positive annotations, clusters were classified as “Unknown” if their annotation score failed to exceed a threshold of 25%. Cell-type-specific differentially expressed genes (DEGs) were identified using FindMarkers implemented in Seurat. Statistical significance was evaluated using the Wilcoxon rank sum test, and gene expression differences with adjusted p-values <0.05 and |log₂ fold change| > 0.25 were considered significant.

### Pseudobulk analysis between PD & control and Δ-specificity scorings

We began with Seurat v5 objects that had per-cell metadata such as cell_type, sample_id (patient), and condition (PD/Control). To avoid pseudoreplication, we conducted analyses at the sample level. We used *scuttle::aggregateAcrossCells* to aggregate raw UMI counts per gene across cells with the same sample_id and condition. Samples containing less than 50 cells (or less than 1% of total cells) were excluded for that cell type. Within each cell_type pseudobulk count matrix, we maintained genes that passed *edgeR::filterByExpr* with the same group factor (PD/Control) to ensure adequate counts in a minimum certain amount of samples per group.

Next, we created an edgeR DGE list, conducted TMM normalization, and fitted a quasi-likelihood negative binomial GLM per gene with design = ∼ condition (PD/Control). Dispersion was assessed using estimateDisp (trended+tagwise), and models were fitted using glmQLFit followed by glmQLFTest for the PD-Control contrast. We utilized ≥2 biological replicates per category for each cell type for calculating statistics. Hits were considered significant when FDR < 0.05 (Benjamini-Hochberg) and |log2FC| > 0.5, unless otherwise specified. We exported complete data (log2FC, QL t, raw P, and FDR) and displayed them using EnhancedVolcano.

To further identify genes that become more or less specific to a cell type in PD, not only globally dysregulated, we calculated Δ-specificity scoring. For each condition, separately and for each cell type. Using similar aggregation by sample method, we built the pseudobulk matrix within each condition. For each cell_type c, construct a one-vs-rest contrast on the pseudobulk counts: model = ∼ group, where group ∈ {c, others}. Fit an edgeR QL GLM and take the estimated log2FC(c vs. rest) as the “specificity score” (g,c) . For every gene g and cell_type c, compute Δ(g, c) as,

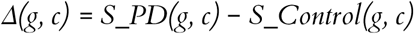

Positive Δ means the gene is more cell-type–specific in PD; negative Δ means loss of specificity in PD (more specific in control).

### Functional categorization of PD-associated gene sets

To summarize PD-associated signatures beyond individual gene lists, we performed a rule-based functional categorization of cell-type-resolved gene sets. We focused on dopaminergic neurons and microglia and used PD-up edgeR-significant genes derived from the sample-aware pseudobulk differential expression framework. Gene symbols were categorized into biologically interpretable functional groups using curated family/prefix pattern matching and gene-class heuristics. Classification was implemented programmatically (Python) using deterministic regular-expression rules anchored to canonical gene family nomenclature (e.g., KMT/KDM/HDAC/KAT/JMJD for chromatin modifiers; CHRM/GRM/GRIA/GRIN/EPH/PTPR/ROBO/UNC5/ADGR for receptors; SCN/CACNA/KCN/RYR/ITPR/ATP2B for excitability/Ca²⁺ handling; NCAM/CADM/CNTN/DSCAM/CDH/COL/TNR for adhesion/ECM/guidance). Genes not confidently assigned to a specific category were labeled Other. Category distributions were summarized per cell type and visualized using pie charts (with “Other” optionally excluded to highlight interpretable functional classes).

### Transcription factor enrichment and regulatory subnetwork analysis

To infer candidate upstream regulators**(Figure 1)** of PD-associated gene programs, we performed TF enrichment analysis using ChEA-KG (ChIP-X Enrichment Analysis Knowledge Graph) (Byrd et al. 2025). TF enrichment was run separately for dopaminergic neuron and microglial gene sets derived from Δ-specificity–prioritized edgeR-significant PD-up genes. ChEA-KG returns (i) a ranked list of enriched TFs and (ii) an enriched TF subnetwork representing directed relationships among enriched TFs. Subnetworks were visualized using ChEA-KG’s standard conventions: nodes represent enriched TFs (highlighting the query TF where applicable), and edges denote curated regulatory relationships labeled as activation or inhibition. Because ChEA-KG integrates multiple evidence channels, we additionally reported the platform’s evidence support summary for enriched TF–target associations, which aggregates support across sources such as coexpression resources and ChIP-seq–derived repositories. ChEA-KG results were treated as hypothesis-generating regulatory context rather than direct causal evidence in the study cohort.

**Figure1:**
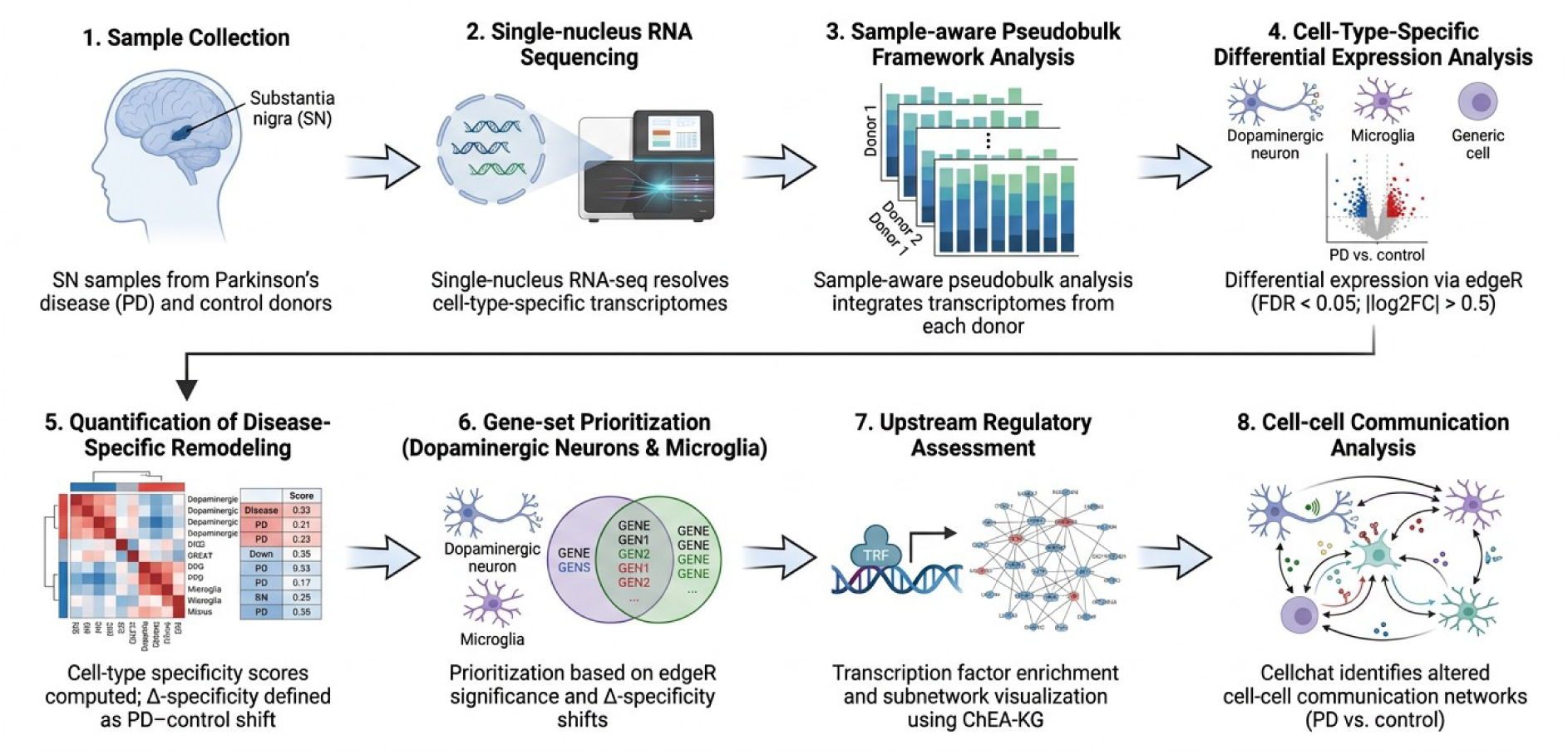
Overview of the study workflow

### Cell-cell communication analysis

Cell-cell communication analysis was conducted using CellChat (v1.6.1) (Jin et al. 2021) to infer the intercellular network between distinct neuronal cells and other cell populations. The CellChat object was created with the “createCellChat” function by utilizing cell-type annotations. The CellChat human database was used for identifying ligand-receptor interactions, and interactions involving fewer than ten cells were filtered out. Communication strength and network centrality were computed at both the interaction and pathway levels, and visualized through network-based approaches. The overview of the study methodology flowchart was created using FigureLabs tool and is provided in **Figure 1**.

## 3. Results

### 3.1 Single-cell profiling of PD and control samples reveal conserved identities with glial remodeling

Single-cell transcriptomic profiling of control and PD samples revealed a consistent set of major neural and non-neural cell populations in both conditions, including dopaminergic neurons, mature neurons, GABAergic neurons, astrocytes, oligodendrocyte precursor cells (OPCs), oligodendrocytes, microglia, and endothelial cells (**Figure 2A**). However, presence of immune-associated cells and radial glial cells in PD samples confirms the disease development (**Figure 2B**). UMAP visualization indicated strong differentiation of these populations based on transcriptional identity, featuring comparable cluster architecture in the control and PD datasets, suggesting that essential cellular identities are preserved regardless of disease status.

**Figure. 2.**
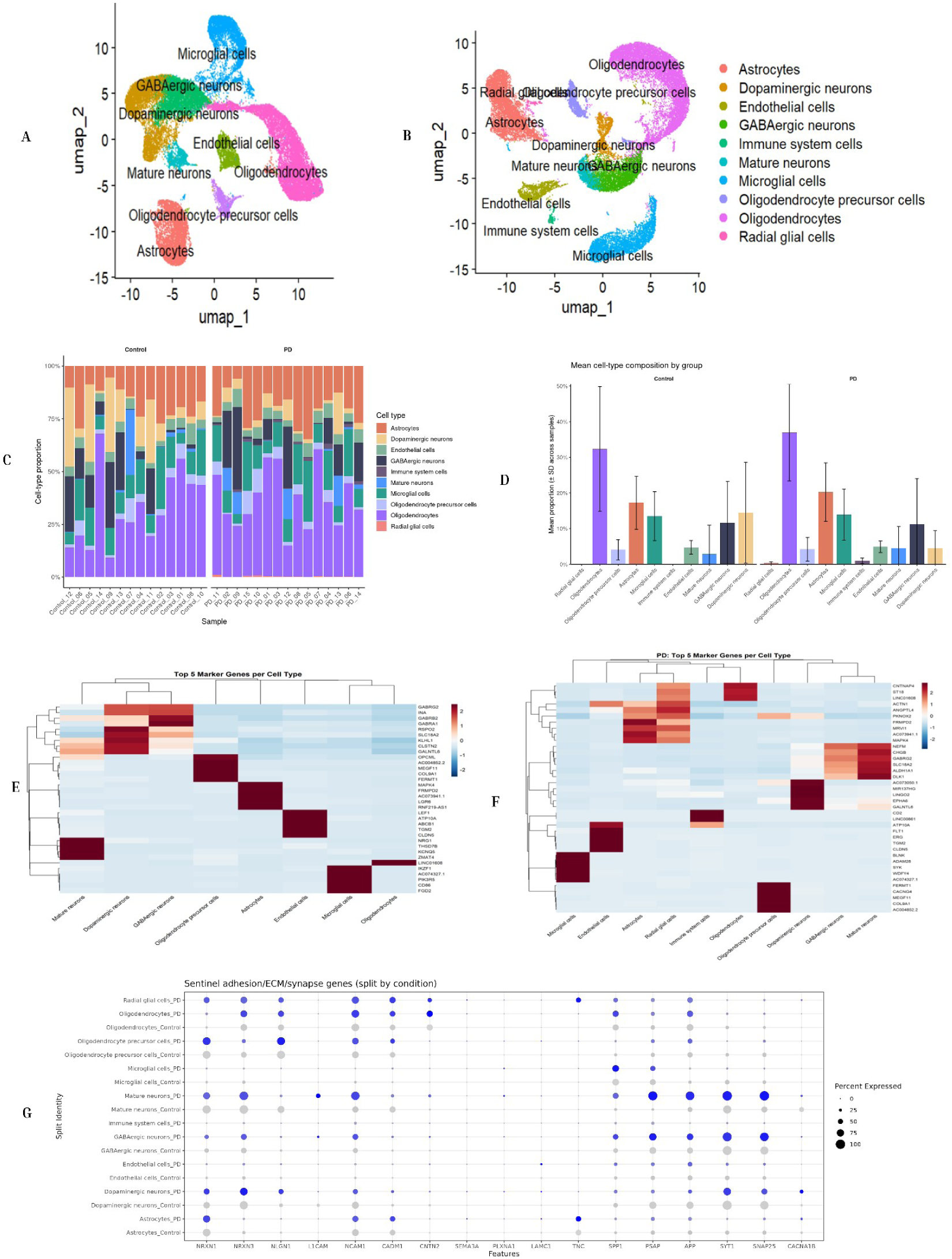
Single nucleus RNA sequencing (SnRNA seq) uncovered cell-type-specific clustering and PD-associated transcriptomic reprogramming in distinct cell subtypes. UMAP plots show annotated clustered in snRNA-seq in **(A)** control and **(B)** PD samples based on sequencing data.**(C)** Relative proportions of neuronal and non-neuronal cell types in control and PD groups . (**D**) ±SD across different samples compared distribution of distinct cell types, depicting significant variations in cell population dynamics in disease compared to healthy controls. Heatmaps show the top five marker genes for each major cell type identified by snRNA-seq in **(E)** control and **(F)** PD samples. (**G**) Dot plot illustrates expression of genes associated with synapsis across cell clusters split by condition. The size represents the percentage of cells within each cluster expressing the gene, while color intensity corresponds to the average expression level.

Although cellular identities were constant, there were significant variations in cell-type composition between control and PD samples. Per-sample composition analysis indicated inter-sample variation in both groups; nevertheless, PD samples were consistently enriched in glial populations, particularly oligodendrocytes, astrocytes, and microglial cells (**Figure 2C**). Group-level analysis revealed an overall rise in the mean proportion of these glial and immune-associated cell types in Parkinson’s disease, accompanied by a relative decrease in neuronal populations, including dopaminergic and GABAergic neurons (**Figure 2D**). These changes point to disease-related modification of the cellular landscape rather than widespread loss or gain of specific cell types except DA neurons.

We calculated the differentially expressed genes (DEGs) across each cell type in both PD and control and identified top marker genes, visualized as heatmaps **(Figure 2E and 2F).** In the control group, dopaminergic neurons demonstrated strong expression of canonical markers (*ALDH1A1 and SLC18A2*), deciphering intact dopaminergic metabolism and vesicular transport. However, in PD, these identity marker genes were significantly reduced along with the emergence of stress-response genes *SYTII* and *HSPA5.* Similarly, others neuronal cell types i.e. GABAergic and mature neurons in the control group revealed upregulation of synaptic and cytoskeletal genes (*NEFM, GABRG2*), but in PD, neuronal marker expression was broadly decreased. Moreover, several neuronal genes were aberrantly observed in glial clusters, suggesting transcriptional reprogramming in PD. Conversely, non-neuronal cell populations also demonstrated shift in expression patterns in disease condition with microglial upregulation of immune effector genes including *CD68, TREM2* and *SYK*. Astrocytes displayed a reactive phenotype in PD enriched in complement (C3) and inflammatory genes (GFAP, VIM). Oligodendrocytes cell subpopulations exhibited decreased myelin-genes and increased stress response markers. Further, Endothelial cells indicated expression of vascular inflammation and barrier disruption genes (*VCAM1, ICAM1 and MMP9).* Moreover, PD samples also contained prominent immune clusters (*CD3D*, *CD79A*, S100A8/A9) and glial markers that were absent in control.

To evaluate alterations in synaptic and structural integrity, we plotted a dot plot to analyze the expression of sentinel adhesion and extracellular matrix (ECM) genes **(Figure 2G).** This analysis demonstrated a shift in gene expression frequency and intensity in PD samples as compared to controls. Synaptic markers such as *SYT1,* and *SNAP25* showed higher expression in neuronal cells, with noticeable increase in the percentage of expressing cells in disease stage. However, adhesion molecules such as NCAM1 and NRXN3 revealed distinct expression patterns across glial and neuronal subpopulations, suggesting a greater reorganization of the cellular environment in PD.

These findings show that Parkinson’s disease is characterized by the maintenance of essential cell-type identities while also undergoing significant cell-type composition change, mostly due to glial and immunological population increase and transcriptional activation. The persistence of canonical marker gene expression across situations shows stable cellular identities, but alterations in abundance and intensity imply changes in disease-associated glial state. This glial remodeling serves as a molecular framework for further understanding pathway dysregulation and altered intercellular communication in Parkinson’s disease.

### 3.2 Pseudobulk differential expression highlights glial remodeling and adhesion/ECM–synapse programs in PD

Using sample-aware pseudobulk DE (PD vs control) within each defined cell type, glia demonstrated the most significant and most organized transcriptional modifications, while neuronal populations undergo parallel, but more targeted, changes (**Fig. 3A-D**). In microglia (**3A**), PD is found to be associated with higher expression of an activation/lipid-handling signature (e.g., APOC1, MAF, CLIC2, TMEM65) and decreased expression of a set of signaling/regulatory genes. This pattern is consistent with microglial state remodeling (Choudhury et al. 2022; Isik et al. 2023; Mirarchi, Albi, and Arcuri 2024), and it confirms the glial-remodeling aspect because the PD-related signal is localized in genes typically linked with immune activation, lipid metabolism, and stress adaptation(Mirarchi et al. 2024).

**Figure. 3.**
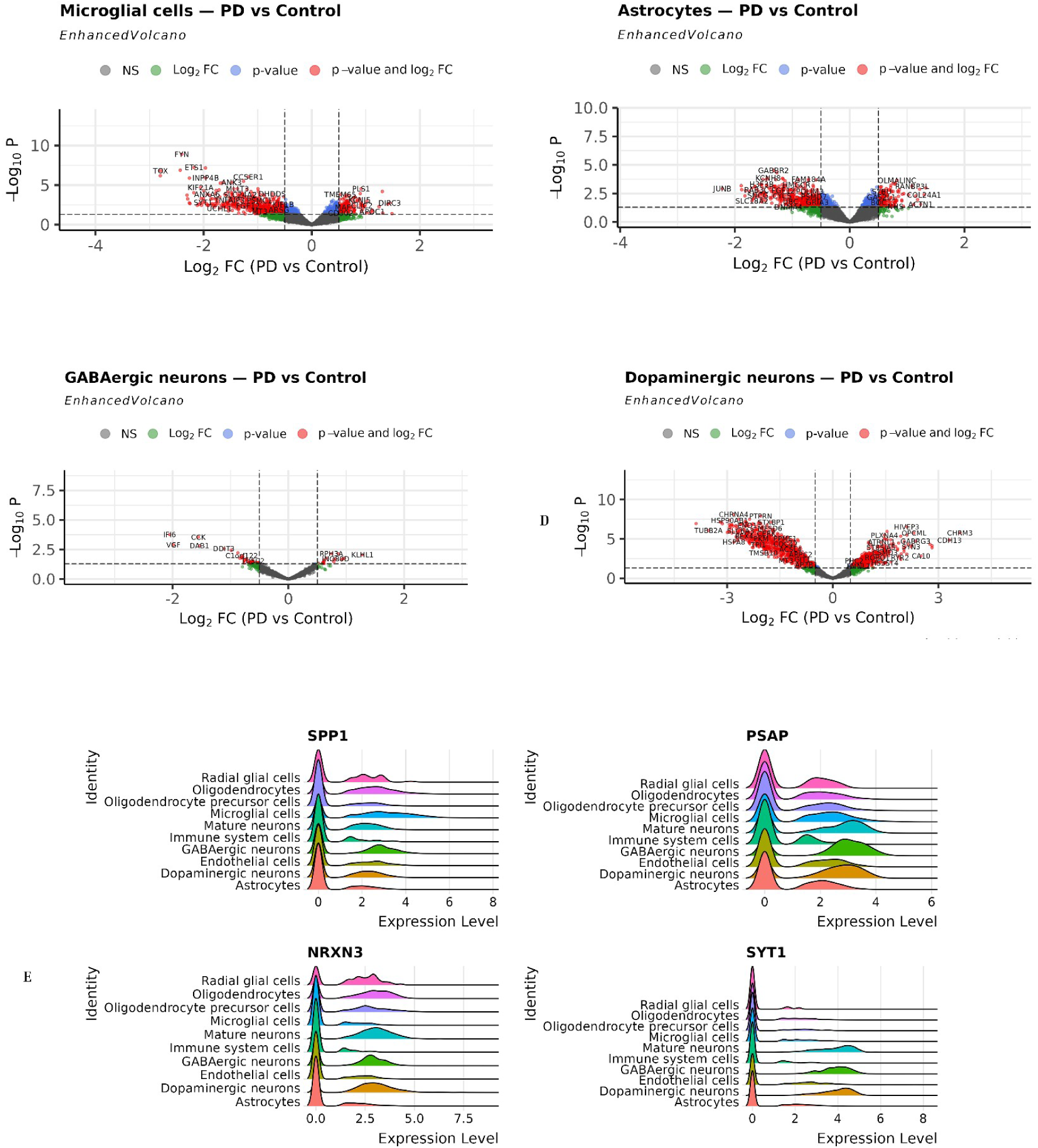
Differential expression of genes in distinct cell types in PD. Volcano plots revealing the differentially expressed genes in 4 different cell types including **(A)** Microglial cells, **(B)** Astrocytes, **(C)** GABAergic neurons, and **(D)** Dopaminergic neurons between PD samples and controls. Y-axis represents − log10_P_ values and X-axis denotes log_2FC. Up-regulated (Right) and down-regulated DEGs (left) in red, non-significant (NS) in gray, with a threshold adjusted p-value ≤ 0.05 and |log2 (FoldChange)| ≥ 0.5. **(E)** Ridge plotS displaying expression distribution of *SPP1, PSAP, NRNX3 and STY1*in neuronal and non-neuronal cells in annotated clusters.

Moreover, Astrocytes (**3B**) show a complementary remodeling signal, with PD-associated increases in structural/ECM-linked components (e.g., COL24A1, ACTN1, and other PD-biased genes identified) and decreases in immediate-early/metabolic or receptor-associated transcripts (e.g., JUNB, HMGCR, GABRB2). Biologically, this suggests a shift toward a more reactive, tissue architecture-influencing astrocyte state, which corresponds to ECM/adhesion remodeling as a mechanistic axis. Neurons exhibit more limited but informative alterations. GABAergic neurons (**3C**) show less significant shifts (fewer labeled extremes), either relative preservation at this resolution or smaller impact sizes detected with pseudobulk. Dopaminergic neurons (**3D**) exhibited broad, highly significant changes, including PD-biased regulation of axon guidance/connectivity genes (e.g., ROBO1, PLXNA4, ATRNL1, ST8SIA3, CSMD3, RYR2, CHRM3), as well as decreased expression of synaptic/secretory or receptor-linked components on the opposite side (e.g., CHRNA4, PTPRN, STXBP1). This is consistent with PD-related dopaminergic vulnerability being accompanied by rewiring/stress responses and changed connection programs.

To further identify cell type specific transcriptional reprogramming, we next plotted a ridge plot based on significantly expressed different genes including *SPP1, PSAP, NRNX3* and *STY1* across annotated cell subpopulations **(Figure 3E)**. *SPP1* expression level was higher predominantly in microglial and oligodendrocytes, with comparatively low expression levels observed in neuronal cell types, consistent with its known association with glial and immune cells activation states (Jakubiak et al. 2024). While *PSAP* showed stronger expression across all cell types with most notable identity in GABAergic neurons. Overall, *PSAP* levels suggested a more ubiquitous functional role. Neuronal genes such as *NRXN3* and *SYT1* were enriched in neuronal populations, specifically in mature and dopaminergic neurons. Microglial and immune system cells have minimal distribution of *NRXN3*, but *SYT1* distributions were lower in non-neuronal cell types. These findings suggest that cellular transition in PD is not limited to dopaminergic neurons while glial populations may be a crucial contributor to PD progression.

### 3.3 Cell-type based gene-level changes show distinct gene-sets across PD and control in neuronal subpopulations

Beyond the differential expression analysis, we calculated Δ-specificity for each gene within its respective cell cluster to evaluate whether PD is affected by cell-type identity reorganization **(Fig. 3)**. In short, we computed a one-vs-rest “specificity score” for each cell type using a sample-aware pseudobulk analysis. Δ-specificity was calculated as the difference in specificity scores between PD and control condition where positive and negative scores indicate gain and loss of cell-type exclusivity in PD, respectively. Across all the annotated cell types, Δ-specificity distributions were centered near zero, indicating that the majority of genes preserve their relative cell-type specificity; however, several subpopulations displayed broader tails, consistent with a subset of genes undergoing substantial PD-associated shifts in cell-type restriction (**Fig. 4A**). Interestingly, hierarchical clustering analysis of Δ-specificity values demonstrated transcriptional coordinated shifts in structured gene modules across distinct cell types **(Fig. 4B)**, supporting the idea that PD implicates systemic reprogramming of cell-type-associated transcriptional networks (Gu et al. 2026.; Martirosyan et al. 2024) rather than an independent gene level change.

**Figure. 4.**
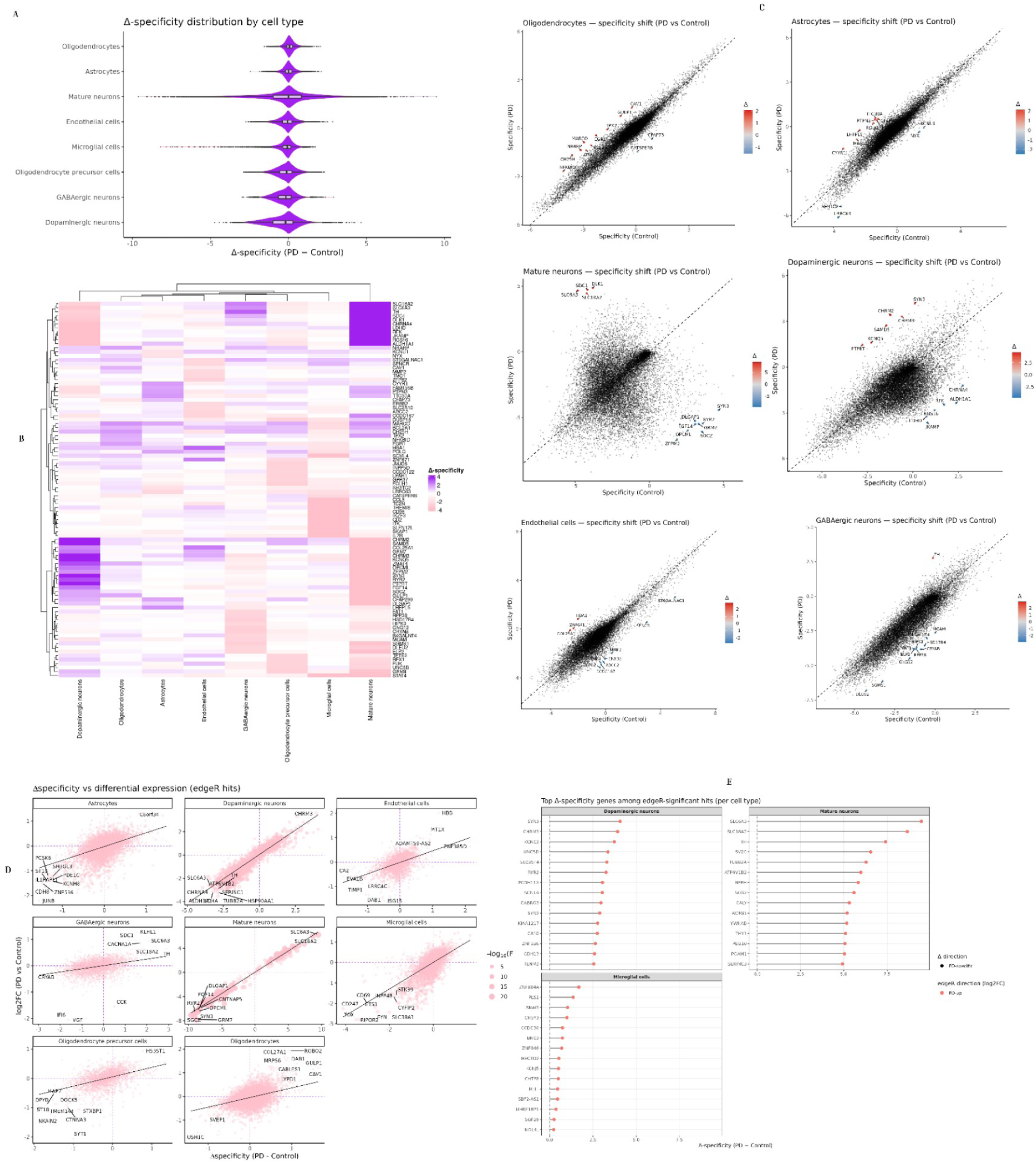
Δ-specificity uncovered predicted potential cell-type-specific candidates in PD progression. **(A)** Violin plot showing Δ-specificity by cell type. Tail length indicates gain or loss of cell-type specificity in PD. **(B)** Heatmaps displaying Δ-specificity for top-ranked genes across cell populations in PD versus Control. Purpule denotes gain specificity in PD while pink denotes loss of specificity. **(C)** scatter plots representing cell-type-specific specificity shifts in 8 different cell subpopulations (oligodendrocytes, astrocytes, mature neurons, dopaminergic neurons, endothelial cells, and GABA neurons) in PD vs Control. Individual dots indicate genes. Blue and red colours denote loss and gain specificity in PD, respectively. **(D)** Biplots comparing log_2FC against Δ-specificity. **(E)** Lollipop charts ranking most prioritized candidates for dopaminergic neurons, astrocytes and microglia. X-axis indicates the magnitude of specificity shift

To further explore gene specificity relationships across each cell type, we compared per-gene specificity in PD relative to Control **(Fig. 4C)**. Across all panels, most genes clustered near the identity line, reflecting relatively stable transcriptomics specificity. Nevertheless, a minority indicated marked deviation and were ranked as strong Δ-specificity candidates. We subsequently relate these identity remodeling findings with expression difference by integrating Δ-specificity with cell-type specific pseudobulk differential expression analysis using edgeR. To better understand this remodeling, we plotted Δ-specificity against log2FC for edgeR hits (**Fig. 5D)** which distinguishes genes having differential expression patterns but with stable specificity from those showing concordant expression dysregulation and specificity redistribution. Therefore, this combination is the strong indicator of cell type specific remodeling in PD rather than a generalized shift. Finally, we selected the edgeR-significant genes having higher Δ-specificity within each cell type **(Fig. 5E**), providing more concise candidate sets for DA, mature neurons and microglia for further mechanistic analysis and validation. Overall, these results support that PD-associated transcriptional shifts in the substantia nigra involve cell-type-specificity reprogramming (Martirosyan et al. 2024; Wang et al. 2024; Zhu et al. 2024) and further combining of Δ-specificity with sample-aware pseudobulk differential expression predict candidate genes implicated in disease-associated remodeling.

**Figure. 5.**
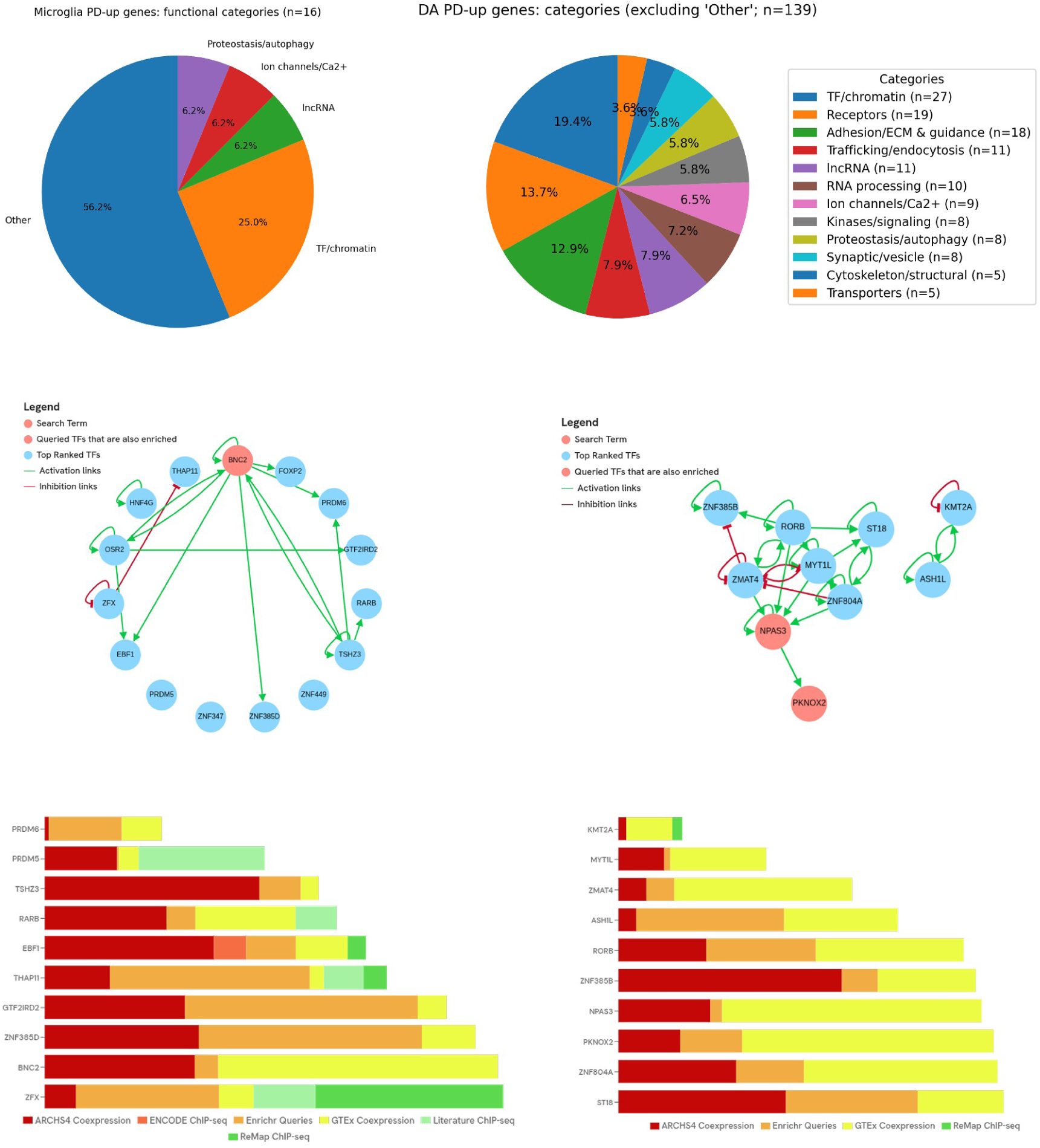
Functional category composition and ChEA-KG TF regulatory subnetworks for PD-up signatures in dopaminergic neurons and microglia. **Top**: pie charts summarizing functional categories for PD-up genes in microglia (n=16) and dopaminergic neurons (excluding “Other” for visualization). **Middle**: ChEA-KG enriched TF subnetworks showing predicted activation (green) and inhibition (red) relationships among enriched TFs; microglia network is centered on BNC2, while the dopaminergic network is centered on NPAS3 and includes ZNF804A and a chromatin-linked component (KMT2A/ASH1L). **Bottom**: evidence supports summary across knowledge sources for enriched TF associations. ChEA-KG results are hypothesis-generating and provide regulatory context for the observed PD-up gene programs.

### 3.4 Candidate gene prioritization highlights dopaminergic connectivity remodeling and microglial state-associated regulatory programs in PD

We next filtered these gene-level findings to a specific set of mechanistically relevant candidates by prioritizing PD-up, edgeR significant genes from dopaminergic neurons and microglia, two cell types central to PD biology. The prioritized PD-up gene set in dopaminergic neurons was strongly enriched for genes involved in neuronal connectivity, Juxtacrine Signaling, receptor responsiveness as well as linking. These key genes include ROBO1, PLXNA4, UNC5D, EFNA5 and CHRM3 **(Table 1).** The dopaminergic neurons reflect consistent patterns in PD-associated transcriptional shifts in connectivity and signaling pathways remodeling which is linked to system level change rather than isolated pathway failure.

**Table 1:** log2FC and FDR are from sample-aware pseudobulk edgeR (FDR < 0.05; |log2FC| > 0.5). Genes were selected from PD-up significant sets in dopaminergic neurons and microglia to provide a compact candidate panel for mechanistic interpretation and downstream validation.

| Cell type | Gene | log<sub>2</sub>FC<br>(PD vs<br>Ctrl) | FDR | Functional<br>bucket<br>(conservative) | Association with PD progression |
| --- | --- | --- | --- | --- | --- |
| Dopaminergic<br>neurons | CHRM3 | 3.609 | 5.10E-05 | Receptor<br>signaling | Strong PD-up receptor-linked<br>signal consistent with altered<br>dopaminergic responsiveness |
|  | PLXNA4 | 1.465 | 7.30E-05 | Axon guidance/<br>connectivity | Guidance receptor candidate<br>supporting dopaminergic wiring<br>remodeling in PD |
|  | UNC5D | 2.758 | 4.10E-04 | Axon guidance/<br>connectivity | Prioritized connectivity cue<br>consistent with altered neuronal<br>contact programs |
|  | EFNA5 | 1.284 | 7.40E-03 | Contact cues/<br>guidance | Contact-dependent signaling gene<br>aligning with adhesion/contact<br>themes |
|  | ROBO1 | 1.264 | 9.30E-03 | Axon guidance/<br>connectivity | Canonical guidance receptor<br>anchoring a PD-associated<br>connectivity program |
| Microglia | PLS1 | 0.914 | 1.18E-02 | Cytoskeleton /<br>motility | Suggests microglial structural<br>remodeling consistent with state<br>transition |
|  | TMEM65 | 0.53 | 2.56E-02 | Mitochondrial-<br>associated | Mitochondrial-linked PD-up<br>candidate consistent with<br>energetic/state remodeling |
|  | MITF | 0.796 | 3.51E-02 | Transcriptional<br>regulator | TF-level candidate that could<br>coordinate a microglial program<br>shift |
|  | HECTD2 | 0.573 | 4.39E-02 | Ubiquitin/<br>proteostasis | Proteostasis-linked remodeling<br>candidate consistent with activation-associated turnover |
|  | KCNJ5 | 0.885 | 4.90E-02 | Ion homeostasis | Ion-channel candidate potentially reflecting functional microglial state changes |
| Shared (DA + Microglia) | ZNF804A | DA: 1.393; MG: 1.308 | DA: 2.38e-02; MG: 1.87e-02 | Regulatory / nuclear | Cross-compartment PD-up candidate (dopaminergic neurons and microglia). |

Among PD-up candidates in microglia, we observed consistent patterns across functional categories underlying the microglia state transitions. These candidates include PLS, MITF, TMEM65, KCNJ5, and HECTD which contribute to cytoskeletal organization and motility, regulatory control, and mitochondrial Ca^2+^ efflux dependent state transition, ionic homeostasis and proteostasis remodeling. Interestingly, the presence of concordantly expressed gene ZNF804A in both cell types may reflect coordinated transcriptional remodeling during PD development **(Table 1)**.

To gather the broader context beyond scattered gene lists, we categorized PD-up genes from dopaminergic neurons and microglia into functional categories. Next, we used ChEA-KG to test which TFs are enriched, giving us insight into both the networks built by these TFs and the evidence backing each association (**Fig. 5**). In microglia, which had 16 PD-up genes, the primary focus is evident: a limited set of functional types explain the changes. Many of these genes belong to TFs as well as chromatin-related functions, accompanied by regulatory and cellular homeostasis roles. This suggests a closely centered, regulatory-heavy microglial program, rather than a dispersed group across many unrelated categories (**Fig5, top-left**). The pattern in dopaminergic neurons looks much more diverse. Setting aside the broad “Other” category, the PD-up genes span a variety of groups: TF/chromatin, receptors, adhesion and ECM guidance, trafficking/endocytosis, RNA processing, ion channel/calcium handling, proteostasis/autophagy, synaptic/vesicle components, kinases and signaling, plus some from cytoskeletal and transporter classes (**Fig5, top-right**). This distribution fits with a remodeling process that juggles variations on the cell surface (receptors and adhesion molecules), shifts in excitability and calcium dynamics, and the mechanism enabling sustained alterations (RNA regulation, trafficking, and proteostasis). These changes aren’t solely focused on one pathway-they impact multiple aspects of neuronal function.

To dig deeper into the regulation behind these PD-up gene programs, we ran TF enrichment analysis through ChEA-KG and mapped the top TF subnetworks for microglia and dopaminergic neurons (**Fig5**, **middle**). In microglia, the TF network centers on BNC2, linking out to several other TFs. This suggests a model where a handful of core TFs coordinate the microglial PD-up program, pointing to a focused, TF-driven regulatory module (**Fig5, middle-left**). Among DA neurons, the situation evolves, the major node is NPAS3, with ZNF804A also woven into the network, with chromatin regulators including KMT2A and ASH1L involved. This leads to a case where transcriptional and epigenetic factors are tightly intertwined, shaping the DA PD-up response (**Fig5, middle-right**). It’s important to keep in mind that these networks are regulatory hypotheses derived from knowledge graphs, not direct proof from these particular samples.

Looking at the ChEA-KG evidence panels (**Fig5, bottom**), we see that the strongest TF connections pull support from a variety of data, from coexpression analyses to ChIP-seq datasets, strengthening the case that these findings aren’t the artifact of a single approach. Taken together, Figure 5 brings the story into focus that DA and microglial PD-up signals sort into distinct, interpretable modules with TF-driven architectures at their core. All this supports the idea that a coordinated dopaminergic–microglial regulatory axis underlies major aspects of PD-related change in the substantia nigra.

Overall, these findings contain a compact, cell-type-specific set of targeted candidates which could be mechanistically interpreted and validated in future and are consistent with the communication network revealed by CellChat.

### 3.5 CellChat reveals disease-associated rewiring of intercellular communication in PD versus control

To further dissect the effect of transcriptional remodeling across distinct cell types in SN which is linked to coordinated changes in intercellular communication network, we predicted altered cell–cell communication patterns between PD and control group using CellChat (v1.6.1) that uses a curated receptor-ligand (LR) interaction database, along with filtering rare interactions. We firstly visualized communication alterations that demonstrated broad engagement of intercellular signaling communication across both neuronal and glial compartment **(Fig. 6A–B**).

**Figure. 6.**
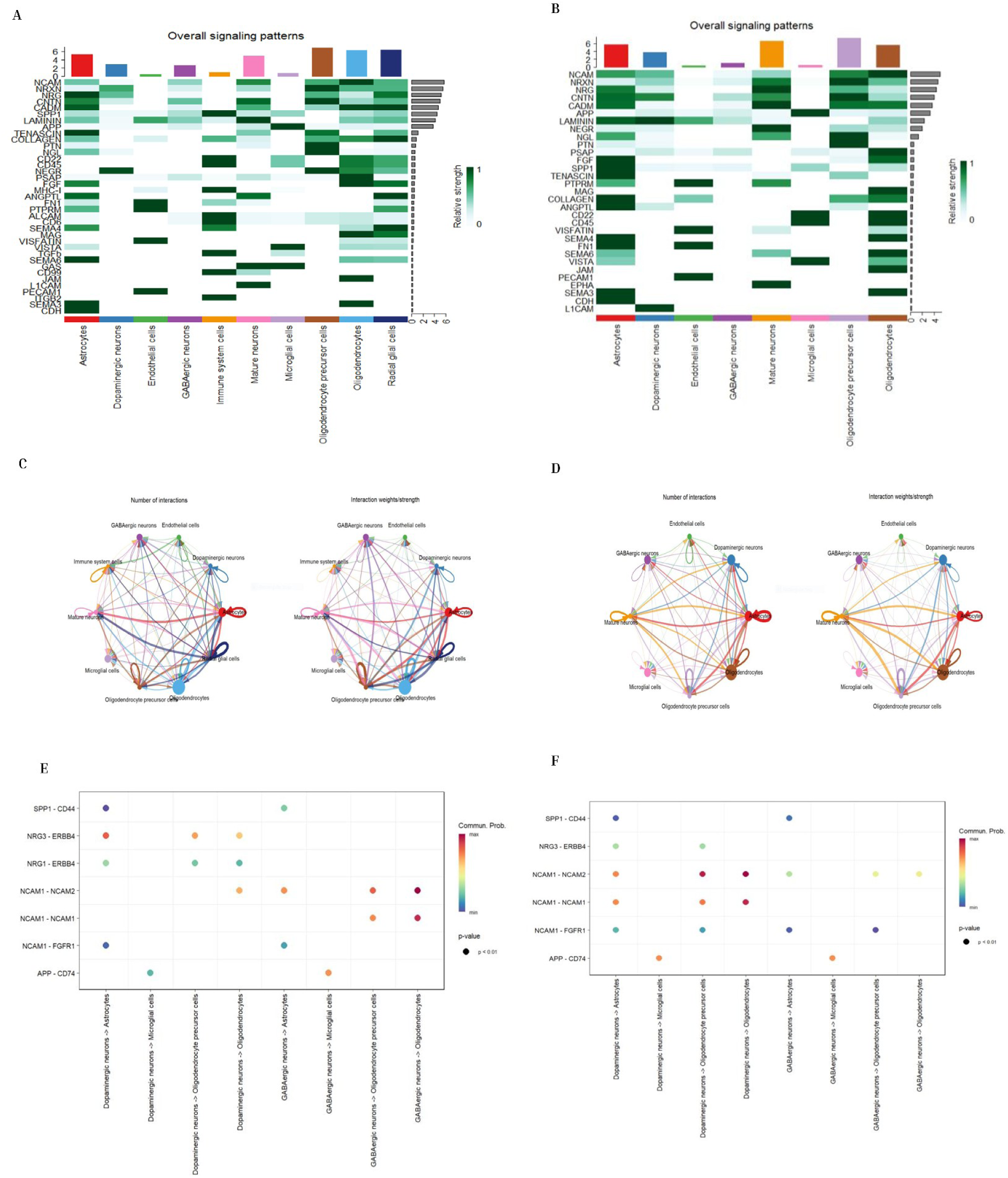
Overview of cell-cell communication predicted by Cellchat in PD and control. Heatmaps summarizing overall singling patterns (consisting of both incoming and outgoing) across each cell type mediated by specific signaling pathways in **(A)** PD and **(B)** control. **(C-D)** Circle plot mapping the number of interactions and interaction strength in each cell population in PD (**left**) and control (**right**). Bubble plot demonstrating the cell-cell communication of prioritized top-ranked ligand-receptors pairs in **(E)** PD (left) and **(F)** control (right). Dot colour indicates the communication probability

In both PD and control samples, the dominant signaling network reflected pathways involved in tissue organization, intracellular connectivity, especially adhesion and ECM associated processes, alongside immune and vascular pathways. Importantly, compared to control **(Fig6B)**, the PD signaling network **(Fig6A**) demonstrated a more widespread distribution of active pathways across more cell types, aligning with expansion of PD-associated intracellular communication network (Huang et al. 2022).

We further explored molecular interactions across cell types using circle plots to infer both the interaction quantity and strength of these interactions. The inferred communication network revealed stronger communication patterns among major neuronal and glial cell clusters in the PD group, reflected by an increased number of interactions and their aggregated strength (**Fig. 6C–D)**. These communication patterns align with previously described PD-associated shifts toward glial cell populations, as supported by pseudobulk analysis indicating glial state remodelling, microglial activation and structural and ECM-related changes in astrocytes. These results indicate that in PD progression, cells undergo not only intrinsic transcriptional changes but also alterations in glia-glia and neuro-glia communication, affecting intercellular communication network

Subsequently, we performed systematic identification of cell communication based on incoming and outgoing signals. The analysis of top-ranked ligand-receptor interactions based on these signals revealed a concise set of genes that consistently observed across neuron-glia crosstalk **(Fig. 6E–F).** Among these prioritized interactions, we found pairs linked to growth factors, cell to cell adhesion and immune-related modules, supporting the notion that PD-associated cellular remodeling alters connectivity matrix and immune signaling. Regarding the dopaminergic neurons pseudobulk analysis results where we found differences in regulation of axon connectivity genes such as ROBO1, PLXNA4, ATRNL1, ST8SIA3, CSMD3, RYR2, CHRM3. However, a decrease in synaptic components (e.g CHRNA4, PTPRN, STXBP1) was observed. Overall, CellChat inference has provided an orthogonal perspective supporting PD-associated neuron–glia interactions in SN.

## 4. Discussion

Parkinson’s disease is defined by degeneration of dopaminergic neurons (Chaudhary, Chaudhary, and Rawat 2025), but SN pathology increasingly appears as a multicellular process in which neuronal vulnerability (Harraz 2023), glial state transitions (Isik et al. 2023; Ma et al. 2025; Mirarchi et al. 2024), and altered trophic/immune signaling co-evolve (Lill et al. 2025; Roodveldt et al. 2024). Large single-nucleus studies of human SN have shown that PD is accompanied by coordinated, cell-type-resolved transcriptional changes and shifts in vulnerable neuronal and glial subpopulations (Gu et al. 2026; Martirosyan et al. 2024; Zhu et al. 2024) . Within this context, our results support a model where PD-associated SN remodeling can be interpreted at three complementary levels: (i) robust, sample-aware pseudobulk differential expression per cell type; (ii) cell-type specificity remodeling (Δ-specificity) indicating identity/program reallocation; and (iii) an inferred communication landscape emphasizing trophic, adhesion/contact, and immune/activation-associated signaling themes Rather than treating these outputs independently, our interpretation focuses on how they converge on a compact set of dopaminergic and microglial candidates. Dopaminergic neuronal loss is recognized as the clinical hallmark of PD (Ben-Shlomo et al. 2024; Chaudhary et al. 2025; Dong-Chen et al. 2023), yet recent evidence showed that surviving nigral neuronal populations undergo adaptive molecular reprogramming, not just maintaining their baseline identity. Evidence from high resolution, single nucleus transcriptomics studies from SN revealed a heterogeneous DA landscape where dopaminergic neurons have distinct subpopulations, and over-representation of vulnerability-associated transcriptional changes in PD such as active stress-adaptive or state-shifted transcriptional programs rather than uniform pool (Martirosyan et al. 2024; Wang et al. 2024). In parallel, mechanistic study on tropic receptor, Neuregulin/(NGR-1)/ErbB4 suggests that modulation of this pathway in PD-associated contexts supports that surviving neurons may activate ErbB4 related trophic signaling as a survival strategy (Depboylu et al. 2012), consistent with a broader “trophic responsiveness” shaping DA state remodeling (Chmielarz and Saarma 2020). These observations are aligned with our PD-up functional architecture including receptor/interface signaling, excitability/Ca²⁺ handling, and RNA/trafficking/proteostasis machinery, supporting a model in which remaining DA neurons represent coordinated transcriptional reprogramming to maintain neuronal function under chronic stress.

Within the glial compartment, microglia state transitions have been reported across multiple studies conducted on SN datasets from PD patients and these transitions are driven by the organized, multi-state programs rather than a single activated phenotype (Liu et al. 2024; Ma et al. 2025; Martirosyan et al. 2024). ScRNA-seq profiling has shown increased microglia and disease-associated glial transcriptional alterations in PD, demonstrating that glial remodeling may be an important feature of PD midbrain (Martirosyan et al. 2024). Evidence from recent snRNA-seq and spatial transcriptomics studies further support that PD-associated glial pathology can be regionally patterned and can colocalize with specific immune features PD brain tissue specimens, suggesting that microglial state programs are sensitive to local cellular niches (Jakubiak et al. 2024; Liu et al. 2024). Our compact microglial PD-up signature showed enrichment for regulatory and state-related genes, supporting the idea of microglia undergoing coordinated transcriptional reprogramming in PD SN. These are linked to previous literature that correlated microglial activation and immune signaling as potential modifiers of PD progression (Appel, Beers, and Henkel 2010; Isik et al. 2023; Mirarchi et al. 2024).

The key feature of this study is that candidate genes mechanisms were not prioritized exclusively from differential expressional analysis but were interpreted via integrated approach, including cell-type-specific remodeling and upstream regulatory structure (Geng et al. 2025; Martirosyan et al. 2024; N M et al. 2024; Pappu et al. 2025). This is closely aligned with the direction of recent SN SnRNA-seq study, which highlights PD-associated remodeling as alterations in cell-type-specific programs and regulatory modules rather than individual DEGs. The ChEA-KG-derived TF subnetworks provide hypothesis-driven regulatory context for these programs by mapping enriched TFs into connected regulatory modules supported by multiple studies i.e. coexpression and ChIP-seq–derived approach, which is suitable for proposing regulatory architectures that may coordinate DA and microglial state shifts without implying direct causal validation. Notably, framing results as regulatory modules are increasingly used to improve interpretability and cross-cohort reproducibility in single-cell disease studies, particularly in heterogeneous tissues such as SN.

Although intercellular communication inference is inherently model-based, it provides a useful systems-level context for interpreting multicellular remodeling when used conservatively. CellChat was explicitly developed to infer and analyze intercellular signaling networks from SnRNA-seq using curated ligand–receptor interaction structure, and its appropriate interpretation is as putative signaling topology rather than direct measurement of ligand–receptor engagement. More broadly, comparative evaluations and reviews of cell–cell communication inference emphasize that results depend on expression thresholds, database content, and the resolution/accuracy of cell-type annotation, underscoring the need to treat CCC outputs as hypothesis-driven, a conceptual framework.

In this study, the value of the communication layer is therefore not to “prove” a specific interaction, but to contextualize DA and microglial candidate modules within a trophic/adhesion/contact/immune-associated signaling landscape that is consistent with the multicellular PD SN environment reported across human and model-system studies.

There are several notable limitations of this study from choosing a study design to the nature of post-mortem SN datasets either single cell or single snRNA transcriptomes. First, CCC and TF-network analysis provided conceptual, regulatory hypotheses rather than causal directionality. This needs to be validated via orthogonal validation either through spatial/protein-level assays or independent cohorts to confirm PD specific pathways and regulators. Second, human SN studies are sensitive to clinical heterogeneity and technical errors, however, sample-aware pseudobulk designs reduce pseudo replication and stabilize inference at the donor level. Moreover, surviving cellular population variability across cohorts can alter effect sizes and the detectability of specific pathways. Finally, cross-study comparisons are further complicated by differences in anatomical sampling (SN vs SNpc), capture chemistry, and annotation granularity. So, careful consideration is required when extending these candidate modules to external datasets.

## Conclusion

Taken together, our results highlight a PD-associated SN remodeling model in which dopaminergic neurons undergo coordinated transcriptional reprogramming consistent with altered disease-responsiveness and interface remodeling. On the other hand, microglia demonstrated enrichment for signatures associated with structured state transitions. TF subnetwork enrichment analysis further supports these programs by providing candidate upstream regulatory modules and the communication network consistent with a multicellular PD SN environment. Overall, this framework shifts focus from descriptive cell-type-specific DE gene lists towards testable, cell-type-resolved regulatory hypotheses that can be validated in future via either independent cohorts or orthogonal spatial modalities.

## Author Contribution

S.N. conceptualized and supervised the study. S.N. and F.Z. performed the bioinformatics analyses and interpreted the data. F.Z. conducted the preliminary processing and CellChat analyses. S.N. performed the integrative downstream analyses, transcription factor network analyses, figure refinement, and manuscript structuring. Both authors contributed to figure generation, manuscript writing, revision, and approved the final version of the manuscript.

## Acknowledgements

The authors acknowledge the researchers and participants of the publicly available Parkinson’s disease single-nucleus transcriptomic study used in this work for generating and sharing the datasets analyzed in this study.

## Data availability

Publicly available single-nucleus RNA sequencing data analyzed in this study was obtained from previously published Parkinson’s disease substantia nigra dataset and their associated repository as mentioned in methodology section. All downstream processed results, gene prioritization tables, and analysis scripts generated during this study are available from the corresponding author upon reasonable request.

